# High sensitivity CDC EUA SARS-CoV-2 kit-based End Point-PCR assay

**DOI:** 10.1101/2020.05.11.20098590

**Authors:** Byron Freire-Paspuel, Patricio Vega-Mariño, Alberto Velez, Marilyn Cruz, Miguel Angel Garcia-Bereguiain

## Abstract

SARS-CoV-2 diagnosis is based on RT-qPCR protocols that limited testing to facilities with Real Time PCR devices and probes supply. Here we described and adapted version of the RT-qPCR CDC protocol where N1, N2 and N3 primers are used for end point PCR and amplicons are visualized on agarose gel with a limit of detection up to 20 viral RNA copies/uL. This protocol would allow to extend SARS-CoV-2 diagnosis to basic molecular biology laboratories with a great impact on surveillance programs at developing countries.

## Background

Since the outbreak of Covid19, the gold standard for SARS-CoV-2 infection diagnosis is RT-qPCR. Several in vitro diagnosis RT-qPCR kits are available on the market SARS-CoV-2 detection. Some of them have received emergency use authorization (EUA) from the U.S. Food & Drug Administration (FDA), like 2019-nCoV CDC EUA from the USA Center for Diseases Control and Prevention (CDC). All RT-qPCR protocols use florescent labelled probes detected on a Real Time PCR devices. For instance, the CDC assay is based on N1, N2 and N3 probes to detect SARS-CoV-2 and RdRP as an RNA extraction quality control (1). These diagnostic tools are basic at molecular biology clinical laboratories on developed countries. However, RT-qPCR is not always accessible for clinical laboratories at developing countries challenging their SARS-CoV-2 testing capacity.

## Objective

This study evaluates the performance of a simple end point PCR protocol for SARS-CoV-2 diagnosis where primers from 2019-nCoV CDC EUA kit (IDT, USA) for viral amplicons N1, N2 and N3 are used to detect up to 20 copies/uL of SARS-CoV-2 on agarose gel electrophoresis. Also RnRP amplicon was visualized for RNA extraction quality control purpose.

## Study design

The end point PCR protocol was calibrated using 2019-nCoV N positive control (IDT, USA) at concentrations from 20 to 20.000 viral copies/uL (Figure 1). Optimal PCR conditions using TaqMan Universal Master Mix (Applied Biosynthesis) and Labnet Multigene (Labnet, USA) thermal cycler were: 50°C 2min; 95°C 10 min; 38 cycles of 3 step: 95°C 30 sec; 56°C 30sec; 72°C 30sec; and final step 72°C 5min. Amplicons were visualized on 3% agarose gels stained with ethidium bromide, and the band sizes were 72 bp, 67 bp, 72 bp for N1, N2 and N3, respectively (Figure 1).

**Figure 1.**
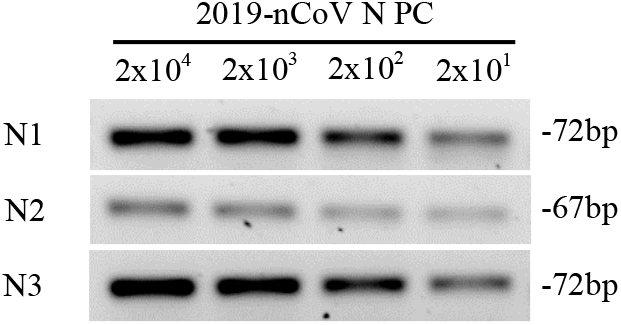
Agarose gel (3%) electrophoresis for “2019-nCoV N PC” positive control for N1, N2 and N3 amplicons.

Thirty-six clinical specimens (nasopharyngeal swabs collected on 0.5mL TE pH 8 buffer) from patients selected as suspicious for SARS-CoV-2 infection were included on this study during the surveillance in Galapagos Islands started on April 8th 2020. Eighteen of the samples were SARS-CoV-2 positive and eighteen were negative, using and adapted version of RT-qPCR CDC protocol (1) with 2019-nCoV CDC EUA kit (IDT, USA), using CFX96 BioRad instrument and PureLink Viral RNA/DNA Mini Kit (Invitrogen, USA) as an alternate RNA extraction method.

## Results

The Ct values and viral loads for the eighteen positive samples are detailed on Table 1, are were from 13.3 to 239000 viral copies/uL. All the positive samples yielded a band on agarose gels for N1, N2, N3 and RdRP, while no band other than for RdRP was obtained with negative samples (Figure 2). The positive sample 972 only yielded a band for N3 (Figure 2) on the first try; but on a new RNA extraction a PCR, samples for N1, N2 and N3 were observed.

**Table 1.**
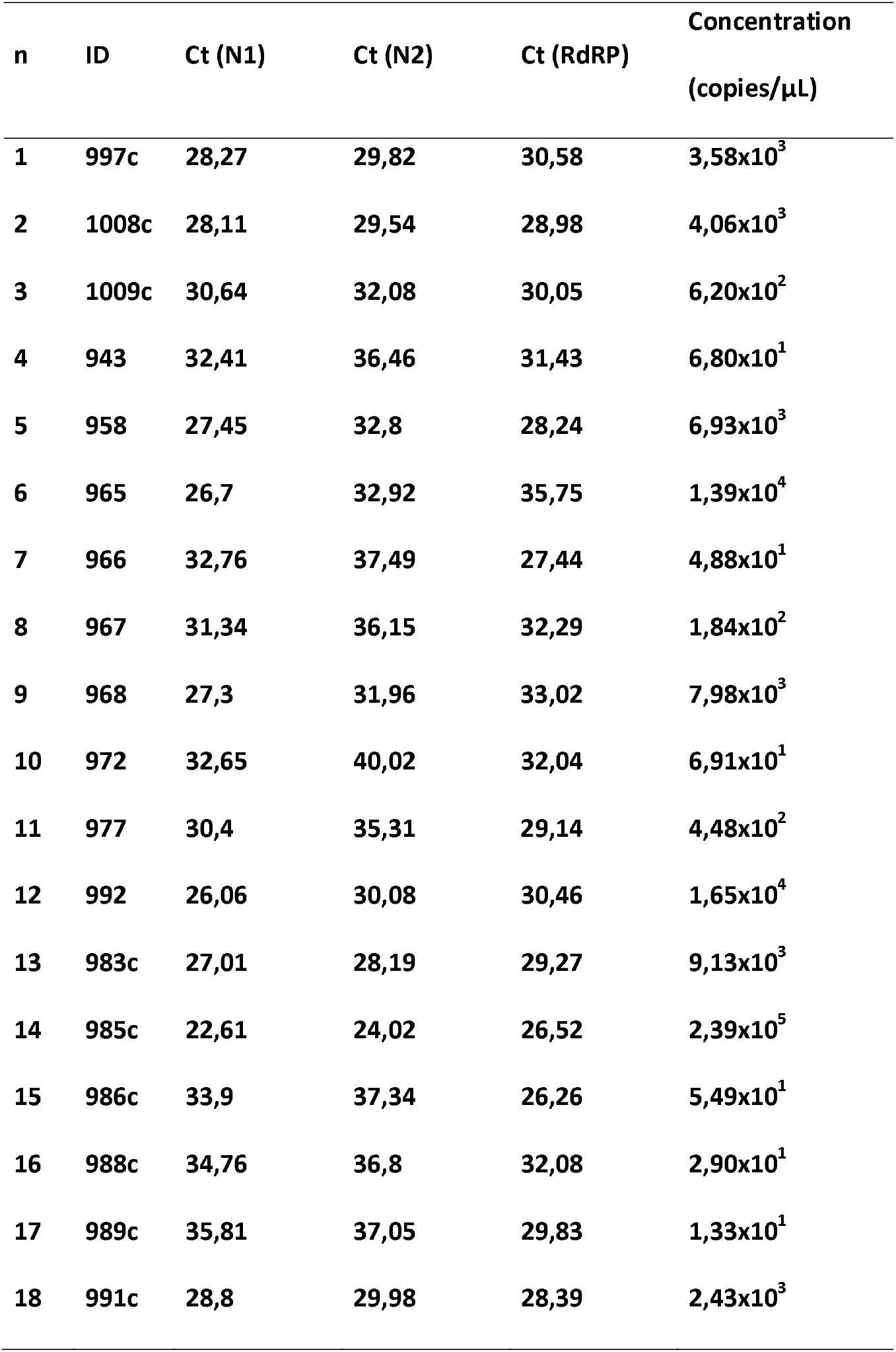
Ct values and viral load (copies/uL) for the eighteen SARS-CoV-2 positive samples included on the end point PCR protocol. Ct values were obtained with the CDC RT-qPCR protocol for N1 and N2 probes.

**Figure 2.**
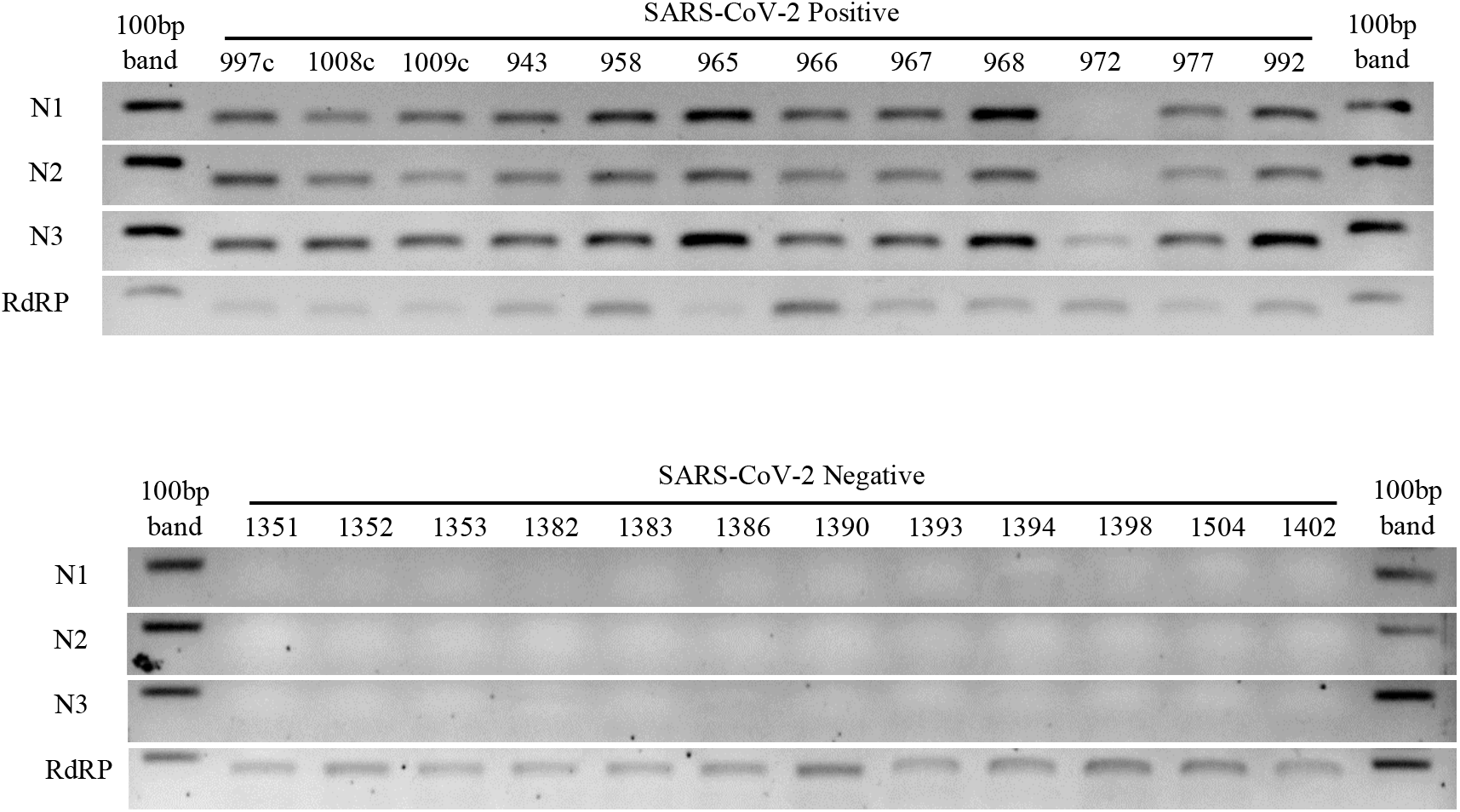
Agarose gel (3%) electrophoresis for SARS-CoV-2 positive and negative samples. N1, N2 and N3 amplicons correspond with viral amplicons for CDC FDA EUA primers set. RdRP amplicons show RNA extraction quality control. Numbers indicates samples code (see supplementary table 1 for Ct values and viral loads). Ten out of eighteen tested positive and negatives are ilustrated.

## Discussion

In summary, we herein described an end point PCR protocol using CDC FDA EUA primers for N1, N2 and N3 viral specific amplicons that allows SARS-CoV-2 detection with 100% specificity and sensitivity as low as least 20 viral RNA copies/uL. As indicated at CDC RT-qPCR protocol, when only N1 or N2 probes are positive, sample must be considered inconclusive and new RNA extraction is recommended (1). Similarly, when only one of the viral amplicons yield a band (like in sample 972), new extraction would allow a more accurate result.

This end point PCR protocol for SARS-CoV-2 diagnosis would be helpful to allow covid 19 surveillance at locations lacking of Real Time PCR devices or without proper funding for probes purchase, with a potential great impact on developing countries.

## Data Availability

Data will be available upon request.

## Acknowledgments

We thank the medical personnel from “Ministerio de Salud Pública” at Galapagos Islands and the staff from the “Agencia de Regulación y Control de la Bioseguridad y Cuarentena para Galápagos” for their support. We specially thank Gabriel Iturralde, Oscar Espinosa and Dr Tannya Lozada from “Dirección General de Investigación de la Universidad de Las Américas” for logistic support to make SARS-CoV-2 diagnosis possible in Galapagos Islands.

## Ethical considerations

All samples have been submitted for routine patient care and diagnostics. Ethical approval for this study was not required since all activities are according to legal provisions defined by the “Comité de Operaciones Especiales Regional de Galápagos” that is leading the Covid19 suerveillance in Galapagos Islands. All data used in the current study was anonymized prior to being obtained by the authors.

## Authorship contribution statement

All authors contributed to study conceptualization, experimental procedures and revision and approval of final version of the manuscript.

Byron Freire-Paspuel and Miguel Angel García Bereguiain analyzed the data and wrote the manuscript.

## Disclaimers

The authors declare no conflict of interest.

## References

1. Interim Guidelines for Collecting, Handling, and Testing Clinical Specimens from Persons for Coronavirus Disease 2019 (COVID-19). Center for Diseases Control and Prevention, USA. https://www.cdc.gov/coronavirus/2019-ncov/lab/guidelines-clinical-specimens.html (last access 04/20/20).

